# Causal relationship between gut microbiota and vulvar cancer: a two-sample bi-directional Mendelian randomization study

**DOI:** 10.1101/2024.08.04.24311470

**Authors:** Jiayan Chen, Peiyan Wang, Changji Xiao, Kalibinuer Kelaimu, Youjie Zeng, Feng Lyu, Xianshu Gao, Xiaomei Li, Jun Hu

## Abstract

**Objective:** Recent investigations have proposed a link between gut microbiomes (GMs) and various cancers, yet the involvement of GMs in vulvar cancer (VC) remains unclear. The objective of this study was to discover the causal association between GMs and VC and identify the GM taxa with potential effect.

**Methods:** Utilizing Mendelian randomization (MR) with genome-wide association study (GWAS) summary statistics, we analyzed 211 GM taxa and 190 VC cases with 167,189 healthy controls. GWAS data for GM taxa were sourced from the MiBioGen consortium, and VC data were acquired from the FinnGen consortium. The main analysis used the inverse-variance weighted (IVW) approach, complemented by weighted median, MR-Egger, weighted mode, and simple mode approaches. Sensitivity analyses included Cochrane’s Q-test, MR-Egger intercept test, MR-PRESSO global test, and leave-one-out analysis.

**Results:** Four nominally significant causal relationships were identified between GM taxa and VC. Class *Betaproteobacteria* [odds ratio (OR)=0.064, 95% confidence interval (CI):0.004-0.946, p=0.045], order *Burkholderiales* [OR=0.074, 95% CI:0.009-0.630, p=0.017], genus *Intestinibacter* [OR=0.073, 95% CI:0.009-0.617, p=0.016], and genus *RuminococcaceaeUCG003* [OR=0.162, 95% CI:0.028-0.938, p=0.042] were linked to a lower chance of VC. The MR-Egger intercept test and MR-PRESSO global test confirmed the lack of horizontal pleiotropy (p>0.05), and leave-one-out analysis indicated result robustness.

**Conclusion:** Our findings highlight four potential causal relationships and specific intestinal flora associated with decreased VC risk, offering insights for VC prevention and treatment.

## 1. Introduction

Vulvar cancer (VC) is a relatively uncommon gynecological malignancy, accounting for approximately 4% of all female genital tract cancers[1] . The worldwide incidence of vulvar cancer stands at 1.2/100,000, with a cumulative lifetime risk of 0.09% [2]. In 2020, a total of 45,240 patients were newly diagnosed of VC and 17,247 deaths were attributed to this disease around the world [2] . A recent epidemiological investigation among 13 high-income countries showed a notable 14% overall increase in the incidence of VC, with uneven distribution across age groups. Women under 60 years experienced a 38% rise in incidence, while no significant change occurred in those aged 60 and older [3].

Squamous cell carcinoma (SCC) is the most common vulvar malignancy, encompassing 80% to 90% of cases[4]. The management of VC poses significant challenges, manifesting in both physical and psychosocial morbidity. Treatment modalities for vulvar cancer span from wide local excision to radical vulvectomy, with or without lymph node biopsy or dissection, and may include radiotherapy with chemo- or immunotherapy for advanced tumors. A noteworthy statistic underscores the challenges, revealing a one-third of mortality rate and a 5-year overall survival of 70.4%[5]. In the absence of specific screening measures, the optimal strategy for reducing the occurrence of VC is the timely treatment of predisposing and preneoplastic lesions related to its oncogenesis. The rareness of the disease has made the study of vulvar squamous cell carcinoma (VSCC) challenging: existing case series tend to be relatively small, and randomised clinical trials are scarce. Thus, further investigations into the pathogenesis and new therapeutic options for vulvar diseases are needed.

The gut microbiomes(GMs) are an ever-changing and intricate system that experiences significant fluctuations influenced by the features of the host [6]. GMs have several important functions, including the metabolization of food ingredients, production of vitamins, defense against infections, preservation of intestinal epithelial barrier, elimination of toxic substances, as well as modulation of inflammatory and immunological reactions[7]. Certain noteworthy associations with microbiome signature associations have been already been identified[8]. However, little is known about the association between vulvar cancer and gut microbiota.

Paucity of available literature and the rarity of the disease limited its further deliberation. MR has evolved as a distinctive research approach akin to randomized controlled trials (RCT) for discovering causal associations between exposures and outcomes[9]. Employing SNPs as IVs, MR leverages the the meiotic genetic randomization, mitigating confounding factors and preempting reverse causation, given the chronological precedence of genetic variants over disease onset[10, 11]. This methodology allows for expedited identification of causal links between exposures and outcomes compared to traditional RCTs. Illustratively, Long et al.’s recent MR study successfully pinpointed GM with potential causal ties to cancers[12]. In this study, we apply MR to extensive GWAS summary statistics of GMs and VC to determine the GM taxa with potential effect, thereby bolstering existing evidence and offering fresh insights into VC prevention and treatment.

## 2. Materials and Methods

### 2.1 Study design

The study’s comprehensive flow chart is depicted in Figure 1. To adhere to the essential assumptions of MR, three criteria were addressed: (i) the IVs exhibit a robust association with exposure factors, (ii) IVs are unrelated to potential confounding variables, and (iii) IVs exclusively influence outcomes through exposure variables[10]. Specifically, the investigation focused on identifying GM taxa with a causal impact on VC through a two-sample MR analysis. The reporting of our findings adhered to the STROBE-MR guidelines[13].

**Figure 1.**
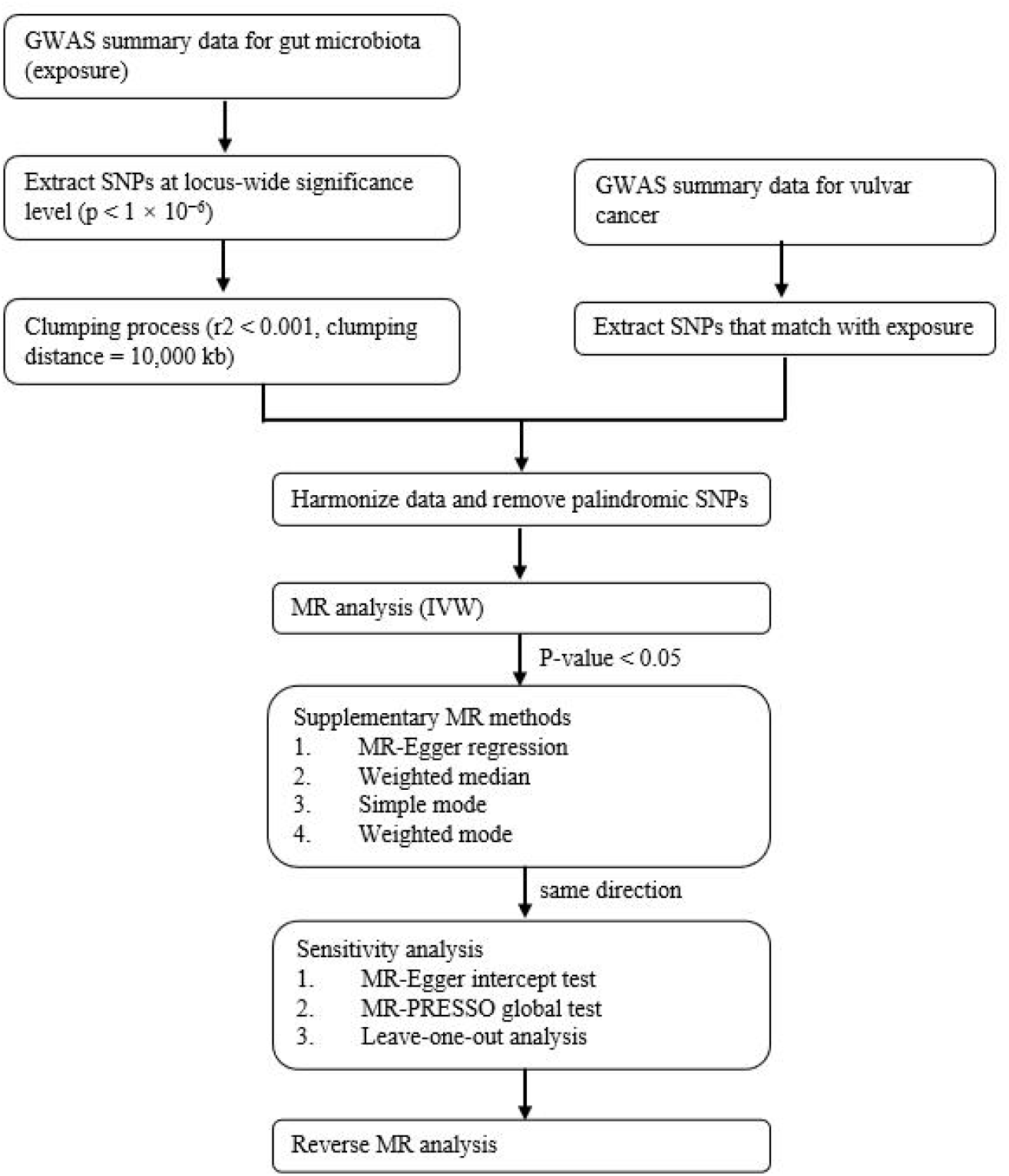
The study design and workflow of the present MR study.

### 2.2 Data sources for the exposure

An investigation conducted by the MiBioGen consortium scrutinized the genotypes of host and 16S fecal microbiome rRNA gene sequencing profiles of 18,340 participants[14]. This GWAS delved into 211 GM taxa spanning from genus to phylum levels, revealing genetic variants related to nine phyla, 16 classes, 20 orders, 35 families, and 131 genera. For those interested, the GWAS summary data of GMs be downloaded from https://mibiogen.gcc.rug.nl/[15–17]

### 2.3 Data sources for the outcome

The GWAS summary data of VC were acquired from the FinnGen consortium R9 release, available at https://r9.finngen.fi/. Comprehensive information regarding the exposure and outcome used in this MR study is presented in Table 1.

**Table 1.**
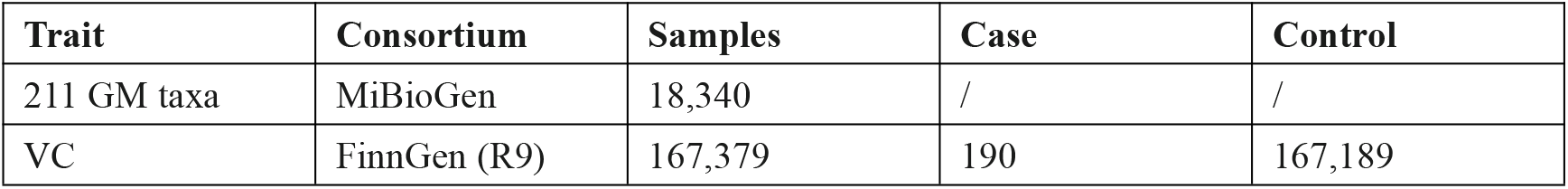
Details of the exposure and outcome.

### 2.4 Instrumental variables

SNPs strongly related to each GM taxon were utilized as IVs. To ensure a sufficiently robust set of IVs, we used a more inclusive threshold (p < 1 × 10^−6^) due to the minimal number obtained under the strict threshold (p < 5 × 10^−8^). For independence, SNPs within a 10,000 kb window size with r^2^ < 0.001 were not adjusted to address linkage disequilibrium (LD). Palindromic SNPs and those not present in the outcome were then removed from the IVs. The level of weak instrumental bias was then assessed using the F-statistic of IVs; an F-statistic >10 denotes the lack of bias as a result of weak IVs [18].

### 2.5 Statistical methods

The main method utilized for causal inference in this MR study was the IVW method. IVW extends the Wald ratio estimator based on Meta-analysis principles[19]. Four additional MR approaches were employed to complement the IVW results when the IVW method demonstrated a causal association (p < 0.05) for each GM taxon: MR-Egger, weighted median, simple mode, and weighted mode [20, 21]. Results of the causal relationship were shown as OR and 95% CI, with a p < 0.05 criterion for significance. FDR correction was implemented for multiple testing, with a threshold of q < 0.05 for various levels. Exposure-outcome pairs were considered to have a causal association only if all MR methods identified the same direction. Sensitivity analyses were performed to assess stability, including the MR-Egger intercept test, MR-PRESSO global test for horizontal pleiotropy[22, 23], and leave-one-out analysis to evaluate result robustness.

To explore the potential causal influence of VC on the significant bacterial genus identified, a reverse MR analysis was carried out, treating VC as the exposure and the reveald causal GM taxa as the outcome. This analysis utilized SNPs linked to VC as IVs.

R software (version 4.2.2) was used for all computations in this investigation. For the MR study, we used the "TwoSampleMR" R package, which is accessible at https://mrcieu.github.io/TwoSampleMR/.

## 3. Results

### 3.1. Details of IVs

In summary, we discovered 16, 26, 26, 54, and 149 SNPs significantly related to gut microbiota on the levels of phylum, class, order, family, and genu, respectively, with a significance threshold of p < 1 × 10^−6^. Furthermore, all IVs exhibited stronger associations with the exposure than with the outcome (pexposure < poutcome), and all F-statistics exceeded 10. For detailed information on the IVs, refer to Supplementary Table S1.

### 3.2. MR analysis

Initially, we conducted a MR analysis to evaluate the causal relationship between 211 GM taxa at different taxonomic levels and VC. The outcomes, assessed by the Inverse Variance Weighted Fixed Effects (IVW-FE) method, indicated that class *Betaproteobacteria* (id: 2867), order *Burkholderiales* (id: 2874), genus *Intestinibacter* (id: 11345), and genus *RuminococcaceaeUCG003* (id: 11361) were linked to a reduced risk of VC (Figure 2). However, post-FDR correction, these associations did not remain statistically significant. Additionally, Cochran’s Q test suggested the lack of heterogeneity in these findings.

**Figure 2.**
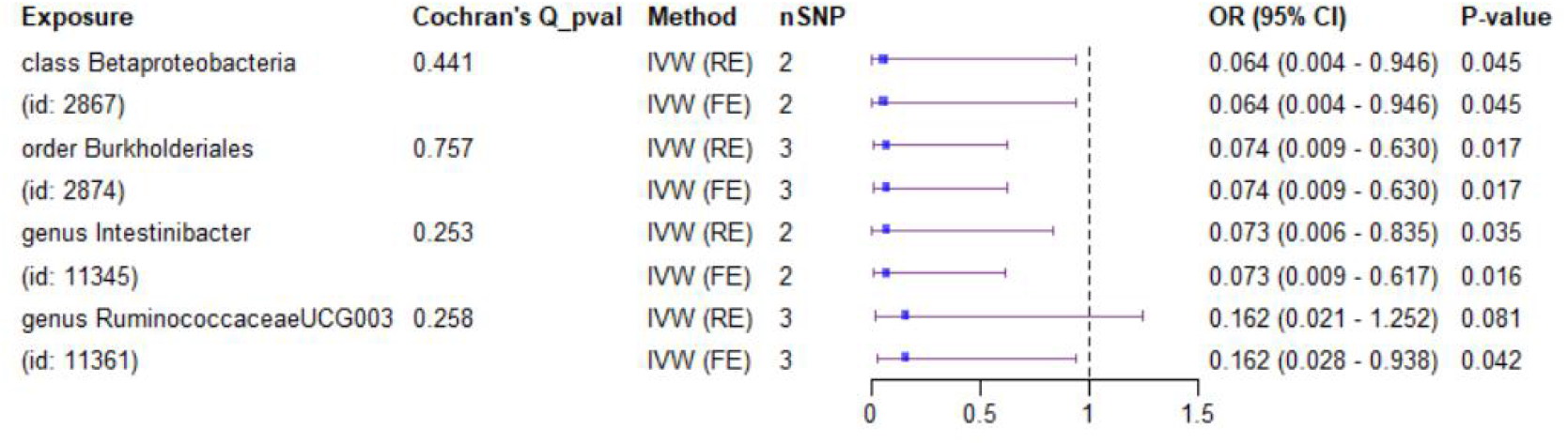
**Forest plot of GM taxa associated with VC identified by IVW_FE method.**

To further verify the causal relationship between these GM taxa and VC, we utilized four more MR techniques: MR-Egger, weighted median, simple mode, and weighted mode (Figure 3). The results were in accordance to the IVW approach consistently.

**Figure 3.**
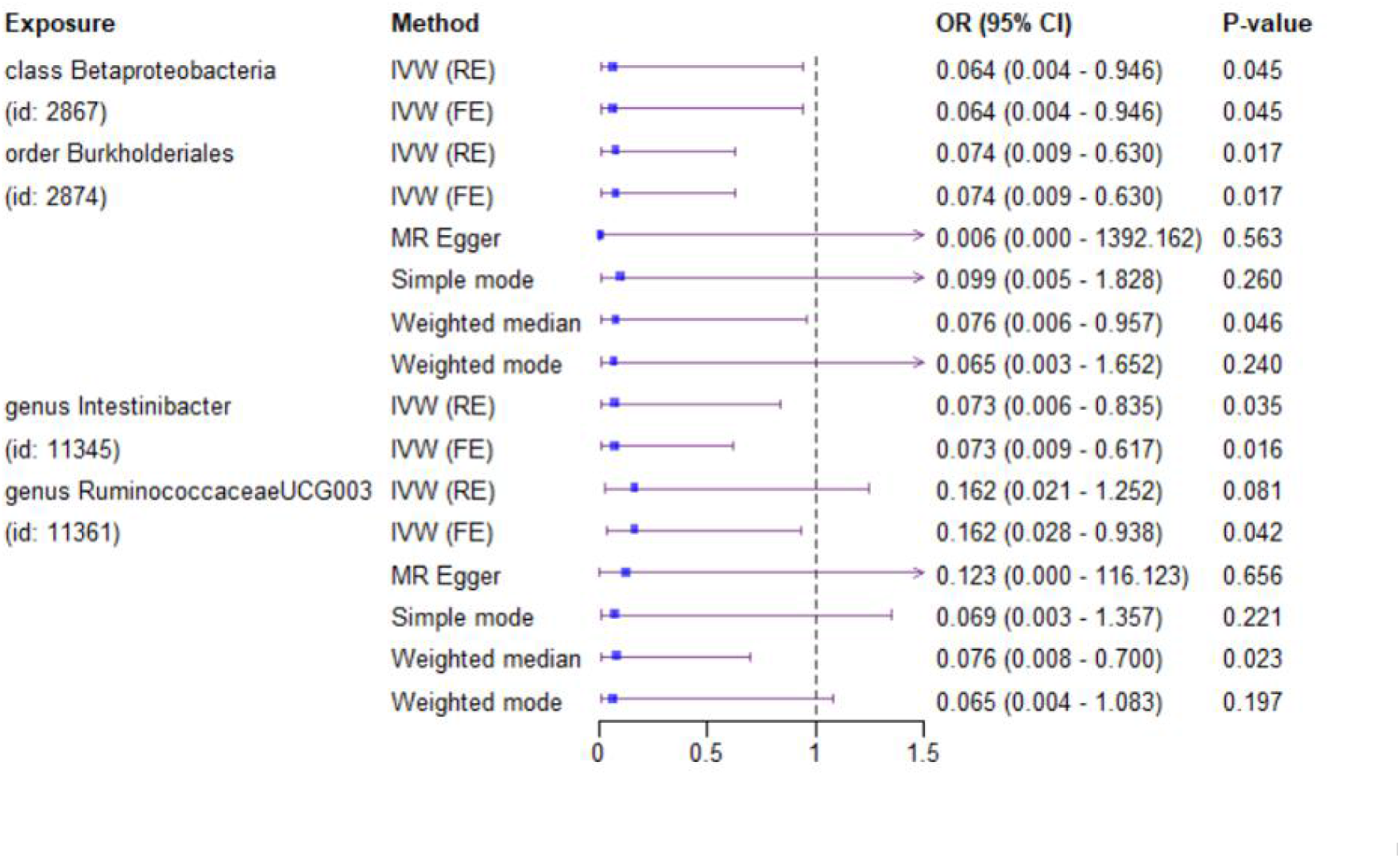
Diverse MR results for 4 GM taxa causally associated with VC.

### 3.3. Sensitivity analysis

The MR-Egger intercept test results revealed no evidence of horizontal pleiotropy (pMR-Egger intercept > 0.05) within the IVs of order *Burkholderiales* (id: 2874) and genus *RuminococcaceaeUCG003* (id: 11361) associated with VC (Supplementary Table S2). Furthermore, the reliability of the MR results was shown by the leave-one-out analysis, since the overall findings remained unchanged to the exclusion of any individual IV (Supplementary Figure S1). Due to the limited number of SNPs for class *Betaproteobacteria* (id: 2867) and genus *Intestinibacter* (id: 11345), the MR-Egger intercept test could not be performed. For the same reason, the MR-PRESSO global test was inapplicable.

### 3.4 Reverse MR analysis

Following a rigorous screening process for IVs, one SNP associated with VC was deemed eligible (Supplementary Table S3). In the reverse MR analysis detailed in Supplementary Table S4, there was no discernible causal effect observed from VC to the identified bacterial features. Given that VC had only one associated SNP, a sensitivity test was not feasible.

## 4. Discussion

Over the past 20 years, extensive research of the composition and oncogenic implications of the gut microbiomes has been conducted, especially in relation to the female reproductive tract tumors. Sims TT et al.’s study examming intestinal bacteria of 42 cervical cancer patients and 46 healthy controls. The results showed that *Prevotella*, *Porphyromonas*, and *Dialister* were in higher abundance in the group of cervical cancer, whereas in the controls, *Acteroides*, *Alistipes*, and *Lachnospiracea* were found of significant enrichment [24]. Li et al reported that the predominant presence of *Proteobacteria*, *Gammaproteobacteria*, *Enterobacteriales*, *Enterobacteriaceae*, and *Shigella* in the endometrial cancer patients[25]. The strong association between the intestinal bacteria, hormone metabolism, and obesity implies a possible contribution from GMs in the progression of hormone-related endometrial cancer. The bacteria may have impact on estrogen enteropathy circulation and associated with hormone-related cancers[26]. Zhou et al. performed a comparative analysis of the microbiome composition in 25 tissue samples from ovarian cancer patients and 25 distal fallopian tube specimens from healthy individuals. The ratio of *Proteobacteria - Firmicutes* exhibited a significant increase in ovarian cancer tissues in comparison to healthy tissue [27]. Notably, a heightened enrichment of *Proteobacteria* was found in the imbalanced intestinal environment, which was considered as a potential indicator for impaired intestinal epithelial function[28, 29]. Current evidence shows that, dysbiosis play a crucial role in inducing changes of the gut barrier, promoting a state of chronic inflammation by activating toll-like receptors. This may result in dysregulated hormones and metabolism. Nevertheless, an comprehensive taxonomic characterization of tumor-related microbiomes is needed, and further mechanistic studies are essential to understand the specific connections between intestinal bacteria and gynecological cancer[30].

Our study undertook a comprehensive assessment of the causal impact of 211 GM taxa on VC. Ultimately, we identified a total of 4 causal relationships, thus highlighting the importance of GMs in VC. Our MR analysis, a pioneering endeavor in this domain, has, for the first time, confirmed that class *Betaproteobacteria* (id. 2867), genus *Intestinibacter* (id. 11345), genus *RuminococcaceaeUCG003* (id. 11361), order *Burkholderiales* (id. 2874) may confer a protective effect on the progression of VC. While the associations observed in this MR study did not attain statistical significance after FDR correction, the potential impact of these gut bacteria may not be disregarded. Alternatively, these findings may indicate a potential GM composition associated with VC that could aid in evaluating the risk of VC. Furthermore, these compositions may also serve as candidate bacteria for investigators to focus on in future functional studies. As an exploratory investigation, our goal was to identify as a range of potential microbial taxa for further research. Therefore, the errors caused by this relatively lenient standard are deemed acceptable to a certain degree.

While there is currently no research indicating a direct connection between class *Betaproteobacteria,* genus *Intestinibacter,* genus *RuminococcaceaeUCG003,* order *Burkholderiales* and VC, these bacteria appear to have a causal or protective effect in malignant diseases. Peters et al. conducted a study on intra tumor and distant normal lung samples from 46 stage II non-small cell lung cancer patients with curative resection[31]. They found that a higher abundance of the classes *Alphaproteobacteria* and *Betaproteobacteria*, as well as the orders *Burkholderiales* and *Neisseriales*, in normal lung tissue was associated with better Recurrence-Free Survival (RFS) [31]. In Han et al’s study, an increased abundance of *Burkholderiales* and a decreased abundance of *Ruminococcaceae* were found to be closely related to colorectal cancer patients with hyperlipoidemia, indicating that alterted gut microorganisms may be involved in the abnormal lipid metabolism, one of the promoters of colorectal cancer[32]. Oyeyemi et al. investigated the microbiome compositions of saliva from the patients with oral squamous cell carcinoma (OSCC), and the result showed *Burkholderiales* were reduced in healthy controls with a corresponding increase in the patients with OSCC [33]. Ma et al ’s MR study revealed an increased abundance of family *Ruminococcaceae*, family *Porphyromonadaceae* and Genus *Bacteroidetes* in the healthy control compared to the samples from hepatocellular carcinoma patients, demonstrating the protective causal associations of these gut microbiomes with liver cancer [34]. However, there was insufficient evidence to conclusively determine the specific role of the identified four gut microbiota in tumor inhibition or anti-tumor immunity of patients. The challenge is to distinguish whether the microbiota itself exerts an inhibitory effect on tumorigenesis(driver), or if the host environment is conductive to certain microflora growth due to genetic factors(passenger). Thus, more investigations are required to reveal the interrelationship among pathologic bacteria, host genes and malignant tumors.

Order *Burkholderiales,* classified under the class *Betaproteobacteria*, are gram-negative bacteria that produce Lipopolysaccharide (LPS). LPS is known to trigger a robust immune response by binding to proteins Toll-like receptor 4 (TLR4) [35]. Although the effect of TLR4 activation on the genesis of tumors may be bidirectional, some immunotherapeutic agents such as Bacillus Calmette–Guérin (BCG), has been demonstrated their role in cancer treatment and received FDA approval [36, 37]. Hosts with favorable responses to tumor therapy or higher anti-tumor immune potential may reveal the enrichment of distinct gut microbiota. *Ruminococcace* has been found to be relatively abundant in the patients with enhanced systemic and antitumor immune responses[38]. Diwakar Davar et al. observed that the resistance to anti-PD-1 immune therapy could be overcome by fecal microbiota transplant, which can reconducted the composition of GM toward the taxa promoting efficacy of anti–PD-1, including *Ruminococcaceae*[39]. Previous research suggests that manipulating and reshaping the GM may serve as an adjuvant anti-cancer option, potentially improving the efficacy of cancer therapy and clinical outcomes[40]. Bacteria and their bioactive compounds producted by bacteria have recently emerged as auspicious anti-tumor therapeutics, with tumor-targeting bacteria providing a unique direction for cancer treatment and prevention [41, 42].

The primary advantages of our study include: (1) In the absence of RCTs, our study expanded the existing research by providing robust evidence regarding the causal association between GM and VC. (2) The analysis involved genetic information of a large sample population, enhancing the validity of these results compared to smaller observational studies. (3) MR analysis helps mitigate confounding factors, allowing for a more robust exploration of causal relationships. The identified causal relationships in our study may provide valuable candidate bacteria for future functional investigations.

However, certain implications and limitations should be considered. Firstly, it is essential to acknowledge that the major subjects of our study were limited to those with European ancestry, and the generalizability of the MR results to other populations warrants further exploration due to population heterogeneity. Secondly, the relaxation of the P-value threshold for selecting instruments and exposures to increase the number of SNPs may pose a higher risk of violating the initial assumption of MR analysis. Nevertheless, the F statistic for all SNPs was over 10, suggesting that no weak SNPs were taken into account in the estimation of MR. Moreover, significant findings underwent rigorous FDR correction to minimize the possibility of false-positive results. Thirdly, while efforts were made to address pleiotropy concerns, the specific biological activities of the utilized SNPs are still unknown at present. However, the consistency of estimates among various MR models and the lack of horizontal pleiotropy in the sensitivity analyses provide reassurance. Fourthly, due to the low incidence of VC, only 190 cases of the disease were included in this study, which is lower than most other MR studies. Thus, we may not only miss other microbiota associated with VC, but also fail to explore the role of GM on different subgroup of this malignancy (such as pathological subtype, age and tobacco usage, etc).

## 5. Conclusions

This MR study establish a foundation of exploring the causal relationship between the gut microbiota and VC. In this study, several intestinal bacteria were identified to have a potential role in reducing the incidence of VC, which may be used for preventing and treating VC. More observational study and basic research are required to verify these findings. In conclusion, this MR study provides insight to the possible causal impact of gut microbiota in the oncogenesis and development of vulvar cancer. These findings indicate that it may be a promising direction of vulvar cancer treatment to modulate gut microbiota and regulate intestinal microenvironment. Further investigation is needed to understand the fundamental mechanism.

## Declarations

The authors declare that the study was conducted in the absence of any commercial or financial relationships that may be considered as a potential conflict of interest.

## Author contributions

JC, PW, XL and JH designed the research. PW collected and analyzed the data. JC, PW, and JH drafted the manuscript. CX and KK helped with data sorting and reviewing. YZ, XG and FL offered technical advices. JH and XL supervised the study and revised the manuscript. Each author made contributions to the paper and accepted the submitted version.

## Ethical Statement

This study used de-identified data accessible to public, which is from participant studies approved by an ethical standards committee. Thus, no additional separate ethical approval was needed for this study.

## Data availability

This investigation examined publicly available datasets. The following links can be used to access this data: https://r9.finngen.fi, https://mibiogen.gcc.rug.nl.

## Data Availability

All data produced in the present work are contained in the manuscript

## Acknowledgements

The authors express gratitude to all the participants of GWAS cohorts involved in the this work and the researchers of the MiBioGen, FinnGen for their kindly sharing the GWAS summary statistics.

## References

[1] A.B. Olawaiye, M.A. Cuello, L.J. Rogers, Cancer of the vulva: 2021 update, International journal of gynaecology and obstetrics: the official organ of the International Federation of Gynaecology and Obstetrics 155 Suppl 1(Suppl 1) (2021) 7-18.

[2] H. Sung, J. Ferlay, R.L. Siegel, M. Laversanne, I. Soerjomataram, A. Jemal, F. Bray, Global Cancer Statistics 2020: GLOBOCAN Estimates of Incidence and Mortality Worldwide for 36 Cancers in 185 Countries, CA: a cancer journal for clinicians 71(3) (2021) 209–249.

[3] Y.J. Kang, M. Smith, E. Barlow, K. Coffey, N. Hacker, K. Canfell, Vulvar cancer in high-income countries: Increasing burden of disease, International journal of cancer 141(11) (2017) 2174–2186.

[4] L.J. Rogers, M.A. Cuello, Cancer of the vulva, International journal of gynaecology and obstetrics: the official organ of the International Federation of Gynaecology and Obstetrics 143 Suppl 2 (2018) 4–13.

[5] F. Bray, J. Ferlay, I. Soerjomataram, R.L. Siegel, L.A. Torre, A. Jemal, Global cancer statistics 2018: GLOBOCAN estimates of incidence and mortality worldwide for 36 cancers in 185 countries, CA: a cancer journal for clinicians 68(6) (2018) 394–424.

[6] V. Lazar, L.M. Ditu, G.G. Pircalabioru, I. Gheorghe, C. Curutiu, A.M. Holban, A. Picu, L. Petcu, M.C. Chifiriuc, Aspects of Gut Microbiota and Immune System Interactions in Infectious Diseases, Immunopathology, and Cancer, Frontiers in immunology 9 (2018) 1830.

[7] E. Thursby, N. Juge, Introduction to the human gut microbiota, The Biochemical journal 474(11) (2017) 1823–1836.

[8] M. Wahid, S.A. Dar, A. Jawed, R.K. Mandal, N. Akhter, S. Khan, F. Khan, S. Jogaiah, A.K. Rai, R. Rattan, Microbes in gynecologic cancers: Causes or consequences and therapeutic potential, Seminars in cancer biology 86(Pt 2) (2022) 1179–1189.

[9] S.A. Swanson, H. Tiemeier, M.A. Ikram, M.A. Hernán, Nature as a Trialist?: Deconstructing the Analogy Between Mendelian Randomization and Randomized Trials, Epidemiology (Cambridge, Mass.) 28(5) (2017) 653–659.

[10] S. Burgess, D.S. Small, S.G. Thompson, A review of instrumental variable estimators for Mendelian randomization, Stat Methods Med Res 26(5) (2017) 2333–2355.

[11] D.A. Lawlor, R.M. Harbord, N.J. Timpson, G.D. Lowe, A. Rumley, T.R. Gaunt, I. Baker, J.W. Yarnell, M. Kivimäki, M. Kumari, P.E. Norman, K. Jamrozik, G.J. Hankey, O.P. Almeida, L. Flicker, N. Warrington, M.G. Marmot, Y. Ben-Shlomo, L.J. Palmer, I.N. Day, S. Ebrahim, G.D. Smith, The association of C-reactive protein and CRP genotype with coronary heart disease: findings from five studies with 4,610 cases amongst 18,637 participants, PloS one 3(8) (2008) e3011.

[12] Y. Long, L. Tang, Y. Zhou, S. Zhao, H. Zhu, Causal relationship between gut microbiota and cancers: a two-sample Mendelian randomisation study, BMC medicine 21(1) (2023) 66.

[13] V.W. Skrivankova, R.C. Richmond, B.A.R. Woolf, J. Yarmolinsky, N.M. Davies, S.A. Swanson, T.J. VanderWeele, J.P.T. Higgins, N.J. Timpson, N. Dimou, C. Langenberg, R.M. Golub, E.W. Loder, V. Gallo, A. Tybjaerg-Hansen, G. Davey Smith, M. Egger, J.B. Richards, Strengthening the Reporting of Observational Studies in Epidemiology Using Mendelian Randomization: The STROBE-MR Statement, Jama 326(16) (2021) 1614–1621.

[14] A. Kurilshikov, C. Medina-Gomez, R. Bacigalupe, D. Radjabzadeh, J. Wang, A. Demirkan, C.I. Le Roy, J.A. Raygoza Garay, C.T. Finnicum, X. Liu, Large-scale association analyses identify host factors influencing human gut microbiome composition, Nature genetics 53(2) (2021) 156–165.

[15] M.A. Swertz, R.C. Jansen, Beyond standardization: dynamic software infrastructures for systems biology, Nature Reviews Genetics 8(3) (2007) 235–243.

[16] M.A. Swertz, M. Dijkstra, T. Adamusiak, J.K. van der Velde, A. Kanterakis, E.T. Roos, J. Lops, G.A. Thorisson, D. Arends, G. Byelas, The MOLGENIS toolkit: rapid prototyping of biosoftware at the push of a button, BMC bioinformatics, BioMed Central, 2010, pp. 1–9.

[17] K.J. van der Velde, F. Imhann, B. Charbon, C. Pang, D. van Enckevort, M. Slofstra, R. Barbieri, R. Alberts, D. Hendriksen, F. Kelpin, MOLGENIS research: advanced bioinformatics data software for non-bioinformaticians, Bioinformatics 35(6) (2019) 1076–1078.

[18] B.L. Pierce, H. Ahsan, T.J. VanderWeele, Power and instrument strength requirements for Mendelian randomization studies using multiple genetic variants, International journal of epidemiology 40(3) (2011) 740–752.

[19] P. Pagoni, N.L. Dimou, N. Murphy, E. Stergiakouli, Using Mendelian randomisation to assess causality in observational studies, BMJ Ment Health 22(2) (2019) 67–71.

[20] J. Bowden, G. Davey Smith, P.C. Haycock, S. Burgess, Consistent estimation in Mendelian randomization with some invalid instruments using a weighted median estimator, Genetic epidemiology 40(4) (2016) 304–314.

[21] S. Burgess, S.G. Thompson, Interpreting findings from Mendelian randomization using the MR-Egger method, European journal of epidemiology 32 (2017) 377–389.

[22] J.M. Rees, A.M. Wood, S. Burgess, Extending the MR-Egger method for multivariable Mendelian randomization to correct for both measured and unmeasured pleiotropy, Statistics in medicine 36(29) (2017) 4705–4718.

[23] M. Verbanck, C.-Y. Chen, B. Neale, R. Do, Detection of widespread horizontal pleiotropy in causal relationships inferred from Mendelian randomization between complex traits and diseases, Nature genetics 50(5) (2018) 693–698.

[24] T.T. Sims, L.E. Colbert, J. Zheng, A.Y. Delgado Medrano, K.L. Hoffman, L. Ramondetta, A. Jazaeri, A. Jhingran, K.M. Schmeler, C.R. Daniel, A. Klopp, Gut microbial diversity and genus-level differences identified in cervical cancer patients versus healthy controls, Gynecologic oncology 155(2) (2019) 237–244.

[25] Y. Li, G. Liu, R. Gong, Y. Xi, Gut Microbiome Dysbiosis in Patients with Endometrial Cancer vs. Healthy Controls Based on 16S rRNA Gene Sequencing, Current microbiology 80(8) (2023) 239.

[26] J.M. Baker, L. Al-Nakkash, M.M. Herbst-Kralovetz, Estrogen-gut microbiome axis: Physiological and clinical implications, Maturitas 103 (2017) 45–53.

[27] B. Zhou, C. Sun, J. Huang, M. Xia, E. Guo, N. Li, H. Lu, W. Shan, Y. Wu, Y. Li, X. Xu, D. Weng, L. Meng, J. Hu, Q. Gao, D. Ma, G. Chen, The biodiversity Composition of Microbiome in Ovarian Carcinoma Patients, Scientific reports 9(1) (2019) 1691.

[28] N.R. Shin, T.W. Whon, J.W. Bae, Proteobacteria: microbial signature of dysbiosis in gut microbiota, Trends in biotechnology 33(9) (2015) 496–503.

[29] Y. Litvak, M.X. Byndloss, R.M. Tsolis, A.J. Bäumler, Dysbiotic Proteobacteria expansion: a microbial signature of epithelial dysfunction, Current opinion in microbiology 39 (2017) 1–6.

[30] F. Borella, A.R. Carosso, S. Cosma, M. Preti, G. Collemi, P. Cassoni, L. Bertero, C. Benedetto, Gut Microbiota and Gynecological Cancers: A Summary of Pathogenetic Mechanisms and Future Directions, ACS infectious diseases 7(5) (2021) 987–1009.

[31] B.A. Peters, H.I. Pass, R.D. Burk, X. Xue, C. Goparaju, C.C. Sollecito, E. Grassi, L.N. Segal, J.J. Tsay, R.B. Hayes, J. Ahn, The lung microbiome, peripheral gene expression, and recurrence-free survival after resection of stage II non-small cell lung cancer, Genome medicine 14(1) (2022) 121.

[32] S. Han, Y. Pan, X. Yang, M. Da, Q. Wei, Y. Gao, Q. Qi, L. Ru, Intestinal microorganisms involved in colorectal cancer complicated with dyslipidosis, Cancer biology & therapy 20(1) (2019) 81–89.

[33] B.F. Oyeyemi, U.S. Kaur, A. Paramraj, Chintamani, R. Tandon, A. Kumar, N.S. Bhavesh, Microbiome analysis of saliva from oral squamous cell carcinoma (OSCC) patients and tobacco abusers with potential biomarkers for oral cancer screening, Heliyon 9(11) (2023) e21773.

[34] J. Ma, J. Li, C. Jin, J. Yang, C. Zheng, K. Chen, Y. Xie, Y. Yang, Z. Bo, J. Wang, Q. Su, J. Wang, G. Chen, Y. Wang, Association of gut microbiome and primary liver cancer: A two-sample Mendelian randomization and case-control study, Liver international : official journal of the International Association for the Study of the Liver 43(1) (2023) 221–233.

[35] C.E. Bryant, D.R. Spring, M. Gangloff, N.J. Gay, The molecular basis of the host response to lipopolysaccharide, Nature reviews. Microbiology 8(1) (2010) 8–14.

[36] M.A. Shetab Boushehri, A. Lamprecht, TLR4-Based Immunotherapeutics in Cancer: A Review of the Achievements and Shortcomings, Molecular pharmaceutics 15(11) (2018) 4777–4800.

[37] E. Vacchelli, L. Galluzzi, A. Eggermont, W.H. Fridman, J. Galon, C. Sautès-Fridman, E. Tartour, L. Zitvogel, G. Kroemer, Trial watch: FDA-approved Toll-like receptor agonists for cancer therapy, Oncoimmunology 1(6) (2012) 894–907.

[38] V. Gopalakrishnan, C.N. Spencer, L. Nezi, A. Reuben, M.C. Andrews, T.V. Karpinets, P.A. Prieto, D. Vicente, K. Hoffman, S.C. Wei, A.P. Cogdill, L. Zhao, C.W. Hudgens, D.S. Hutchinson, T. Manzo, M. Petaccia de Macedo, T. Cotechini, T. Kumar, W.S. Chen, S.M. Reddy, R. Szczepaniak Sloane, J. Galloway-Pena, H. Jiang, P.L. Chen, E.J. Shpall, K. Rezvani, A.M. Alousi, R.F. Chemaly, S. Shelburne, L.M. Vence, P.C. Okhuysen, V.B. Jensen, A.G. Swennes, F. McAllister, E. Marcelo Riquelme Sanchez, Y. Zhang, E. Le Chatelier, L. Zitvogel, N. Pons, J.L. Austin-Breneman, L.E. Haydu, E.M. Burton, J.M. Gardner, E. Sirmans, J. Hu, A.J. Lazar, T. Tsujikawa, A. Diab, H. Tawbi, I.C. Glitza, W.J. Hwu, S.P. Patel, S.E. Woodman, R.N. Amaria, M.A. Davies, J.E. Gershenwald, P. Hwu, J.E. Lee, J. Zhang, L.M. Coussens, Z.A. Cooper, P.A. Futreal, C.R. Daniel, N.J. Ajami, J.F. Petrosino, M.T. Tetzlaff, P. Sharma, J.P. Allison, R.R. Jenq, J.A. Wargo, Gut microbiome modulates response to anti-PD-1 immunotherapy in melanoma patients, Science (New York, N.Y.) 359(6371) (2018) 97–103.

[39] D. Davar, A.K. Dzutsev, J.A. McCulloch, R.R. Rodrigues, J.M. Chauvin, R.M. Morrison, R.N. Deblasio, C. Menna, Q. Ding, O. Pagliano, B. Zidi, S. Zhang, J.H. Badger, M. Vetizou, A.M. Cole, M.R. Fernandes, S. Prescott, R.G.F. Costa, A.K. Balaji, A. Morgun, I. Vujkovic-Cvijin, H. Wang, A.A. Borhani, M.B. Schwartz, H.M. Dubner, S.J. Ernst, A. Rose, Y.G. Najjar, Y. Belkaid, J.M. Kirkwood, G. Trinchieri, H.M. Zarour, Fecal microbiota transplant overcomes resistance to anti-PD-1 therapy in melanoma patients, Science (New York, N.Y.) 371(6529) (2021) 595–602.

[40] W. Li, X. Deng, T. Chen, Exploring the Modulatory Effects of Gut Microbiota in Anti-Cancer Therapy, Frontiers in oncology 11 (2021) 644454.

[41] P. Baindara, S.M. Mandal, Bacteria and bacterial anticancer agents as a promising alternative for cancer therapeutics, Biochimie 177 (2020) 164–189.

[42] E.M. Park, M. Chelvanambi, N. Bhutiani, G. Kroemer, L. Zitvogel, J.A. Wargo, Targeting the gut and tumor microbiota in cancer, Nature medicine 28(4) (2022) 690–703.

