## Supplemental Figures for "Causal relationship between gut microbiota and vulvar cancer: a two-sample bi-directional Mendelian randomization study"

A

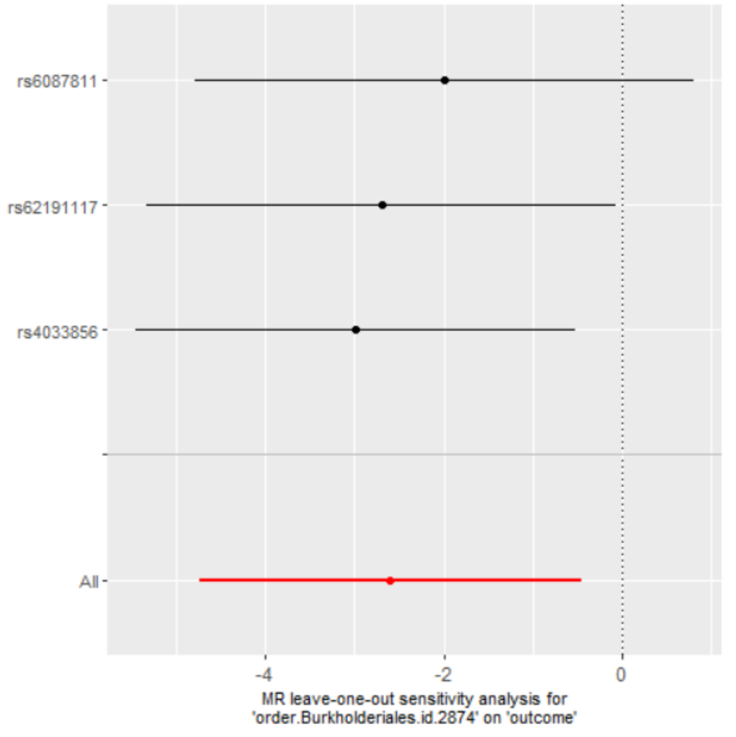


B

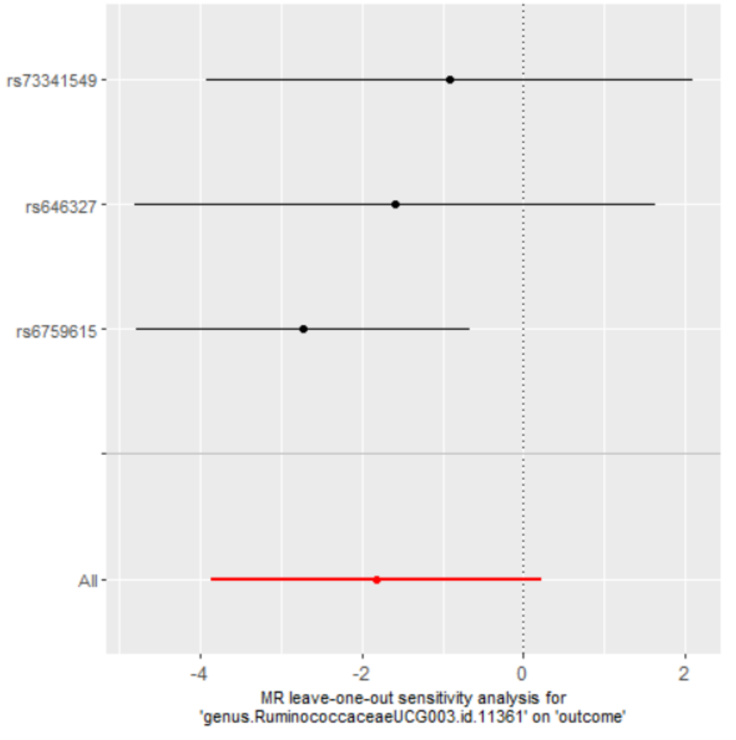


Supplementary Figure S1. Leave-one-out analysis of the causal effect of VC on *Burkholderiales* (A) and *RuminococcaceaeUCG003* (B). Red lines represent estimations from the IVW test. The number of SNPs of *Betaproteobacteria* and *Intestinibacter* is insufficient for leave-one-out analysis.
